# FOLIC ACID AND METHOTREXATE USE AND THEIR ASSOCIATION WITH COVID-19 DIAGNOSIS AND MORTALITY: AN ANALYSIS FROM THE UK BIOBANK

**DOI:** 10.1101/2022.02.10.22270804

**Authors:** Ruth K Topless, Ralph Green, Sarah L. Morgan, Philip C Robinson, Tony R Merriman, Angelo L. Gaffo

**Affiliations:** Department of Biochemistry, University of Otago, Dunedin, New Zealand; Departments of Pathology and Medicine, University of California Davis at Sacramento, Sacramento, California, USA; Division of Clinical Immunology and Rheumatology, The University of Alabama at Birmingham, Birmingham, Alabama, USA; School of Clinical Medicine, University of Queensland, Brisbane, Australia; Birmingham VA Medical Center. Birmingham, Alabama, USA

## Abstract

**Importance:** Folate metabolism is implicated in SARS-CoV-2 infectivity. Medication affecting folate metabolism may influence the risk of COVID-19 diagnosis and outcomes.

**Objective:** to determine if methotrexate (an antifolate) or folic acid prescription were associated with differential risk, for COVID-19 diagnosis or mortality.

**Design, Setting, and Participants:** Case-control analysis of COVID-19 from the population-based UK Biobank (UKBB) cohort. Updated medical information was retrieved on the 13^th^ December 2021. Data from 380,380 UKBB participants with general practice prescription data for 2019 to 2021 were used. Criteria for COVID-19 diagnosis were 1) a positive SARS-CoV-2 test or 2) ICD-10 code for confirmed COVID-19 (U07.1) or probable COVID-19 (U07.2) in hospital records, or death records. By these criteria 26,003 individuals were identified with COVID-19 of whom 820 were known to have died from COVID-19. Logistic regression statistical models were adjusted for age sex, ethnicity, Townsend deprivation index, BMI, smoking status, presence of rheumatoid arthritis, sickle cell disease, use of anticonvulsants, statins and iron supplements.

**Exposures:** Prescription of folic acid and/or methotrexate.

**Main outcomes and measures:** The outcomes of COVID-19 diagnosis and COVID-19 related mortality were analyzed by multivariable logistic regression. The odds ratios from different exposures were compared.

**Results:** Compared with people prescribed neither folic acid nor methotrexate, people prescribed folic acid supplementation had increased risk of diagnosis of COVID-19 (OR 1.51 [1.42 ; 1.61]). The prescription of methotrexate with or without folic acid was not associated with COVID-19 diagnosis (P≥0.18). People prescribed folic acid supplementation had positive association with death after a diagnosis of COVID-19 (OR 2.64 [2.15 ; 3.24]) in a fully adjusted model. The prescription of methotrexate in combination with folic acid was not associated with an increased risk for COVID-19 related death (1.07 [0.57 ; 1.98]).

**Conclusions and Relevance:** We report increased risk for COVID-19 diagnosis and COVID-19-related death for people prescribed folic acid supplementation. Prescription and use of supplemental folic acid may confer increased risk of infection with SARS-CoV-2 and increased risk of death resulting from COVID-19. Our results indicate that methotrexate attenuates an increased risk for COVID-19 diagnosis and death conferred by folic acid.

**Key Points:** *Question:* Does folate supplementation and/or methotrexate use affect the risk COVID-19 diagnosis and COVID-19 associated mortality?

*Findings:* In this epidemiological analysis from the UK Biobank, folic acid supplementation was associated with a 1.5-fold increased risk of COVID-19 diagnosis and a 2.6-fold increased risk of COVID-19 associated mortality. Methotrexate use might attenuate an increased risk for COVID-19 diagnosis and death conferred by folic acid.

*Meaning:* Folic acid supplementation appears to be associated with increased risk for COVID-19 diagnosis and associated mortality while methotrexate use attenuated this risk

## Introduction

Folate, a B-vitamin, carries out critical roles in the transfer of one-carbon units in intermediary metabolism. Folates exist in various forms depending on the one-carbon substituent attached to the parent molecule and are involved in numerous reactions, including the synthesis of methionine from homocysteine and are also utilized in purine and pyrimidine metabolism for DNA and RNA synthesis. The oxidized form, folic acid, is added to fortified foods in the USA and over 80 other countries, including, recently, the UK^1^ to prevent neural tube defect pregnancies and is used in dietary supplements to prevent or treat folate deficiency.^2^ Additionally, folic acid supplementation of up to 5mg daily is often advised during pregnancy and in women of childbearing age and for other medical conditions (sickle cell anemia) ^3^ and during treatment with certain anticonvulsants ^4^.

Methotrexate, a structural analogue of folate has potent antifolate activity and is in widespread use as an antineoplastic agent and as a first-line disease-modifying antirheumatic drug (DMARD) treatment for rheumatoid arthritis (RA).^5^ Folic acid (at doses commonly ranging from 1-2 mg daily) or folinic acid supplementation is often included to lower the toxicity of low-dose methotrexate therapy. ^6,7^

The COVID-19 Global Rheumatology Alliance physician-reported registry has evaluated factors related to death from COVID-19 in individuals with rheumatic diseases.^8^ Compared with those receiving methotrexate monotherapy, use of rituximab (OR 4.0 [95% CI 2.3 ; 7.0]), sulfasalazine (3.6 [1.7 ; 7.8]), azathioprine, cyclophosphamide, cyclosporine, mycophenolate, and tacrolimus (2.2 [1.4 – 3.4]) or no DMARD (2.1 [1.5 – 3.0]) all had higher risks of death from COVID-19.

In order to generate purines SARS-CoV-2 post-transcriptionally remodels host folate metabolism. In an *in vitro* system using African green monkey kidney cells infected with SARS-CoV-2 intracellular glucose and folate were depleted, and this perturbation was sensitive to folate inhibitors such as methotrexate. ^9^ It is therefore plausible that methotrexate therapy for RA could have a beneficial effect on COVID-19 outcomes given its antifolate activity. However, since folic acid is routinely included with methotrexate to prevent methotrexate-related toxicity, such putative beneficial effect of methotrexate on viral proliferation and hence on COVID-19 outcomes may be negated by folic acid supplementation.

The aim of this study was to determine whether the use of methotrexate and folic acid prescription, together or individually, were associated with a lowered or increased risk, respectively, for COVID-19 diagnosis or mortality in a large population based-cohort.

## Participants and Methods

### Data availability

This research was conducted using the UK Biobank Resource (approval number 12611). The UK Biobank is a large resource of volunteers aged 49-86 years of age at recruitment.^10^ Recruitment began in 2006 with follow-up intended for at least 30-years. SARS-CoV-2 test information, ICD-10 hospital codes, death records and general practice prescription information were obtained via the UK Biobank data portal on 13^th^ December 2021. This information covered hospital diagnoses between 18^th^ April 1991 and 30^th^ September 2021, SARS-CoV-2 tests between 13^th^ January 2020 and 18^th^ October 2021, and death records until 12^th^ November 2021. Illustrated in Figure 1 there were 464,306 participants, of whom 4,469 were removed owing to not having a BMI measure or Townsend index score or smoking status and a further 79,457 were removed owing to lack of prescription data. General practice prescription data from 1st of January 2019 through to 27^th^ September 2021, available for 380,380 participants, were used to identify people prescribed methotrexate, folic acid, anticonvulsants (phenytoin, carbamazepine, phenobarbital), iron supplements (ferrous fumarate, ferrous sulfate, ferrous gluconate), and co-prescribed medications. We did not involve patients or the public in the design, or conduct, or reporting, or dissemination plans of our research.

**Figure 1.**
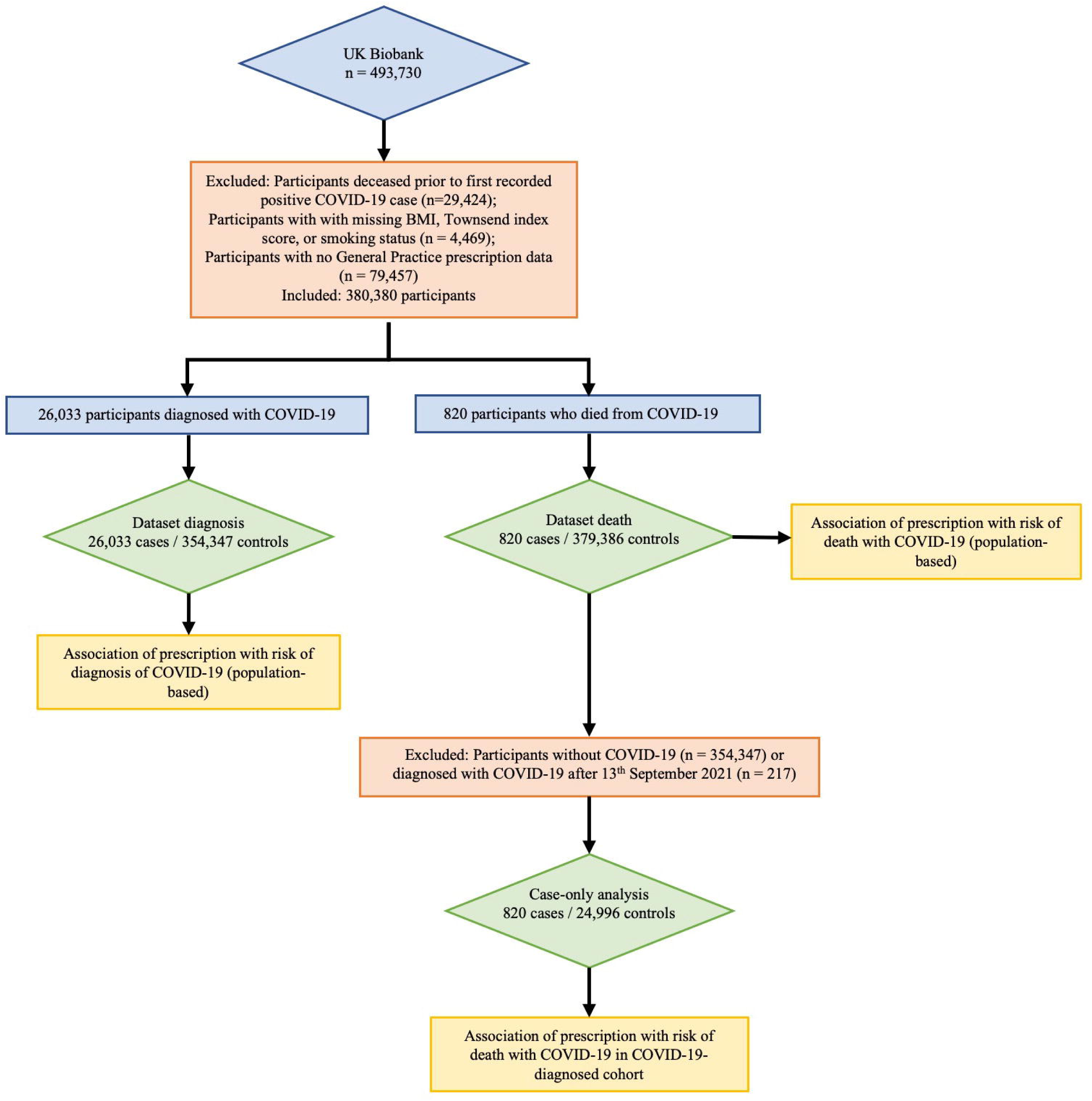
Flow schematic of study design.

### COVID-19 definitions

The criteria for COVID-19 diagnosis were defined as participants with 1) a positive SARS-CoV-2 PCR test and / or 2) ICD-10 code for confirmed COVID-19 (U07.1) or probable COVID-19 (U07.2) in hospital records, or death records. There were 26,033 cases, of whom 820 died with COVID-19. Figure 2 summarizes how cases were diagnosed.

**Figure 2.**
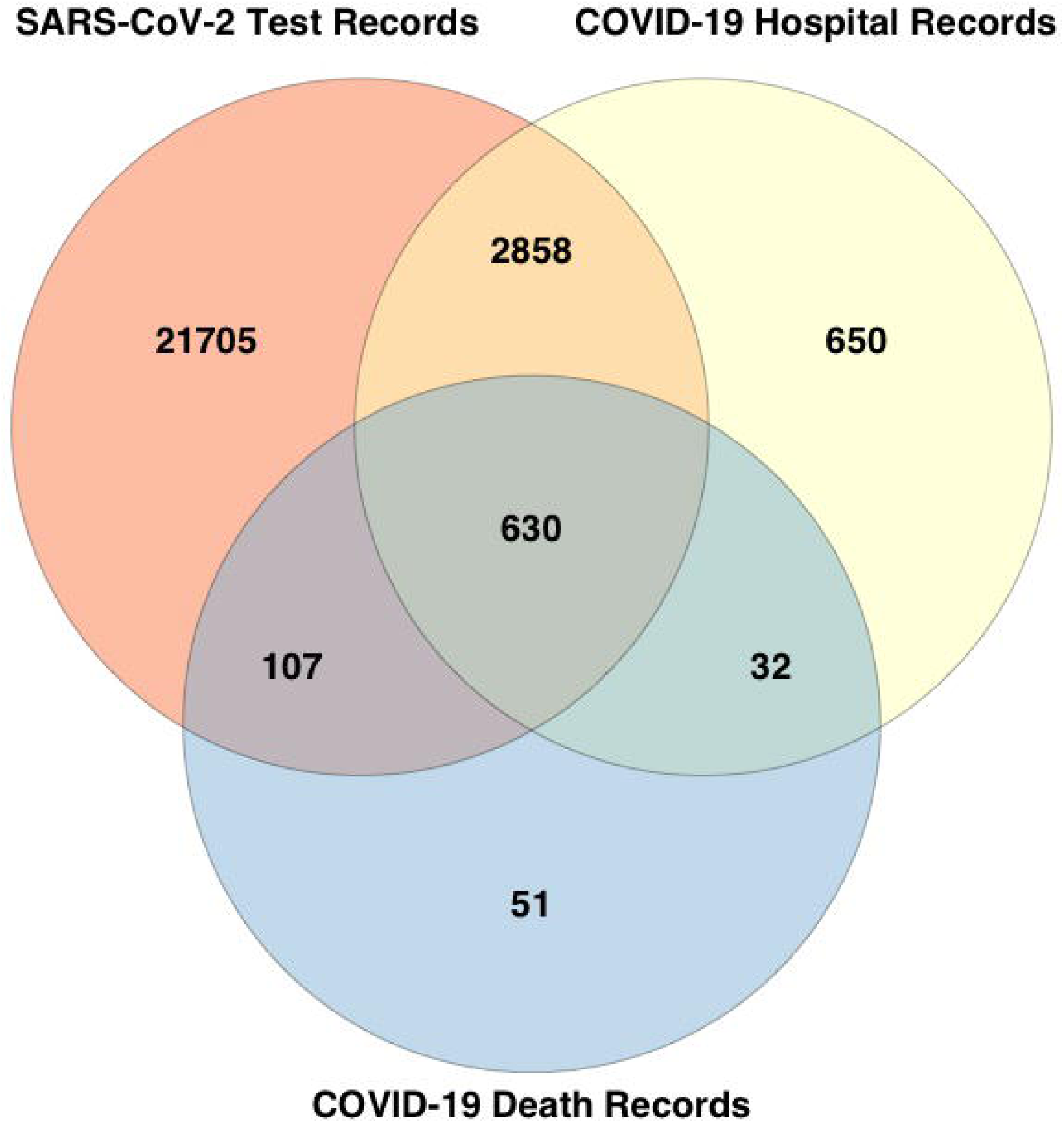
Data sources of COVID-19-diagnosed individuals. Of the 26,033 COVID-19-diagnosed individuals, 25,300 were identified from positive SARS-Cov2 test results (21,705 unique to this group), 4,170 identified from hospital records (650 unique to this group), and 820 identified from death records (51 unique to this group). 217 diagnosed after 13^th^ October 2021 (30 days before the last recorded death) were removed from the case only analysis in Table S4 given the unknown outcome.

### Ethnicity, age and comorbidity data

Self-reported ethnicity was grouped into White British (British, Irish, White, any other White background), Black British (African, White and Black African, Black or Black British, Caribbean, White and Black Caribbean, Any other Black background), Asian British (Asian or Asian British, Chinese, Indian, Pakistani, Bangladeshi, White and Asian, Any other Asian background), and Other (Other ethnic group, Mixed, Any other mixed background, Do not know, Prefer not to answer). Age was calculated for 2020 from year of birth. Age groups used in the analysis were <60 years (n= 69,849), 60-69 years (n= 120,013), 70-74 years (n= 90,627) and >74 years (n= 99,891). The ICD-10 hospital codes used to determine additional comorbidity status were rheumatoid arthritis (M05), and sickle cell disease (D57). Demographic characteristics of the study population are presented in Table 1.

**Table 1.** Study population in the UK Biobank, restricted to those with data on prescriptions

| Demographic | All<br>N= 380,380 | COVID-19 diagnosis<br>N= 26,033 | COVID-19-related death<br>N= 820 |
| --- | --- | --- | --- |
| <60 years of age, n(%) | 69,849 ( 18.36 ) | 7,937 ( 30.49 ) | 26 ( 3.17 ) |
| 60-70 years of age, n(%) | 120,013 ( 31.55 ) | 8,568 ( 32.91 ) | 129 ( 15.73 ) |
| 70-75 years of age, n(%) | 90,627 ( 23.83 ) | 4,569 ( 17.55 ) | 171 ( 20.85 ) |
| >75 years of age, n(%) | 99,891 ( 26.26 ) | 4,959 ( 19.05 ) | 494 ( 60.24 ) |
| Female sex, n(%) | 211,363 ( 55.57 ) | 13,802 ( 53.02 ) | 286 ( 34.88 ) |
| White British, n(%) | 357,620 ( 94.02 ) | 23,807 ( 91.45 ) | 744 ( 90.73 ) |
| Black British, n(%) | 9,826 ( 2.58 ) | 1,021 ( 3.92 ) | 36 ( 4.39 ) |
| Asian British, n(%) | 7,329 ( 1.93 ) | 732 ( 2.81 ) | 25 ( 3.05 ) |
| Other ethnicity, n(%) | 5,605 ( 1.47 ) | 473 ( 1.82 ) | 15 ( 1.83 ) |
| Prescribed methotrexate only, n(%) | 174 ( 0.05 ) | 11 ( 0.04 ) | 0 ( 0 ) |
| Prescribed folic acid only, n(%) | 12,433 ( 3.27 ) | 1,273 ( 4.89 ) | 120 ( 14.63 ) |
| Prescribed methotrexate and folic acid, n(%) | 3,952 ( 1.04 ) | 287 ( 1.10 ) | 11 ( 1.34 ) |
| Prescribed neither methotrexate nor folic acid, n(%) | 363,821 ( 95.65 ) | 24,462 ( 93.97 ) | 689 ( 84.02 ) |
| Rheumatoid arthritis, n(%) | 999 ( 0.26 ) | 97 ( 0.37 ) | 8 ( 0.98 ) |
| Sickle cell disease, n(%) | 517 ( 0.14 ) | 51 ( 0.20 ) | 2 ( 0.24 ) |
| Prescribed anticonvulsant medication, n(%) | 1,642 ( 0.43 ) | 120 ( 0.46 ) | 10 ( 1.22 ) |
| Prescribed statins, n(%) | 156,064 ( 41.03 ) | 10,398 ( 39.94 ) | 521 ( 63.54 ) |
| Prescribed iron supplements, n(%) | 18,471 ( 4.86 ) | 1,661 ( 6.38 ) | 106 ( 12.93 ) |
| BMI, mean(sd) | 27.41 ( 4.76 ) | 28.12 ( 5.03 ) | 30.21 ( 5.79 ) |
| Townsend deprivation index, mean (sd) | -1.35 ( 3.04 ) | -0.9 ( 3.18 ) | -0.28 ( 3.40 ) |
| Never smoked, n(%) | 210,993 ( 55.47 ) | 13,990 ( 53.74 ) | 321 ( 39.15 ) |
| Current smoker, n(%) | 132,222 ( 34.76 ) | 9,262 ( 35.58 ) | 384 ( 46.83 ) |
| Former smoker, n(%) | 37,165 ( 9.77 ) | 2,781 ( 10.68 ) | 115 ( 14.02 ) |

### Statistical analysis

All association analyses were done using R v4.0.2 in RStudio 1.2.5019. Statistical model 1 was adjusted for age group (4 categories), sex, ethnicity, Townsend deprivation index, BMI, smoking status. Model 2 is Model 1 plus adjustment for the presence of rheumatoid arthritis, sickle cell disease (where daily folic acid is prescribed ^3^), prescription of statins, prescription of anticonvulsants (where co-prescription of folic acid often occurs ^11^) and iron supplements (supplementary iron has been associated with poorer outcomes of infectious disease, including COVID-19 ^12,13^). A p < 0.05 threshold indicated nominal evidence for association.

## Results

### Study population

Demographic characteristics of the study population are presented in Table 1. The proportion of those diagnosed with COVID-19 while taking methotrexate was similar to the general study population (1.14% vs 1.09%, respectively) although there was a higher proportion of methotrexate prescriptions in the group that died of COVID-19 (1.34%). There was both a higher proportion of those prescribed folic acid who were diagnosed with COVID-19 (5.99% vs 4.31% in the general population) and those prescribed folic acid in those who died with COVID-19 (15.97% vs 4.31% in the general population). Medications co-prescribed with folic acid were investigated (Table S1). Atorvastatin was co-prescribed 23.21% of the time and Simvastatin 9.49% of the time. Due to these high prescription rates and reports describing an association between statin use and reduced mortality from COVID-19 ^14,15^ we included statins in Model 2.

### Association with a diagnosis of COVID-19

Compared with people prescribed neither folic acid nor methotrexate, individuals prescribed folic acid had significant association with diagnosis of COVID-19 in Model 1 (OR 1.60 [1.50 ; 1.70]) (Table 2). In Model 2, which included a diagnosis of RA, sickle cell disease, and prescription of anticonvulsants or statins or iron supplements, this association was not attenuated (OR 1.51 [1.42 ; 1.61]). The prescription of methotrexate without folic acid was uncommon (only 174 people) and did not show an association with COVID-19 diagnosis in either Model. The prescription of methotrexate in combination with folic acid was associated with an increased risk for a diagnosis of COVID-19 in Model 1 (1.15 [ 1.02 ; 1.30]) but not in Model 2 (1.09 [0.96 ; 1.23]) (Table 2). The risk for COVID-19 diagnosis was associated with similar magnitudes with the prescription of folic acid in men and women in Model 2 (OR 1.50 [1.37 ; 1.64] and 1.52 [1.39 ; 1.65], respectively) (Table S2). The Model 2 sex-specific associations were not statistically significant and of similar magnitudes with methotrexate and with methotrexate combined with folic acid.

**Table 2.**
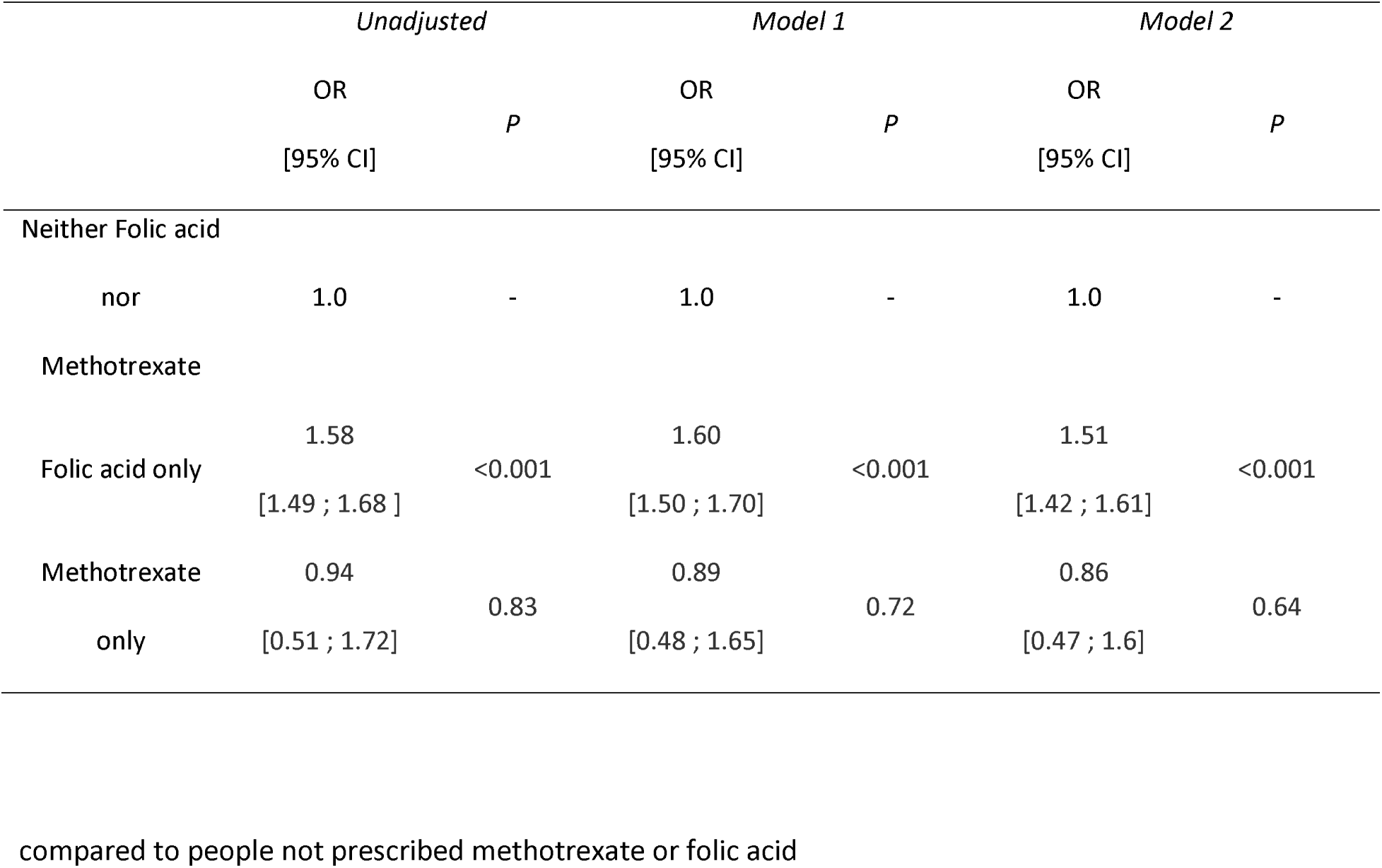
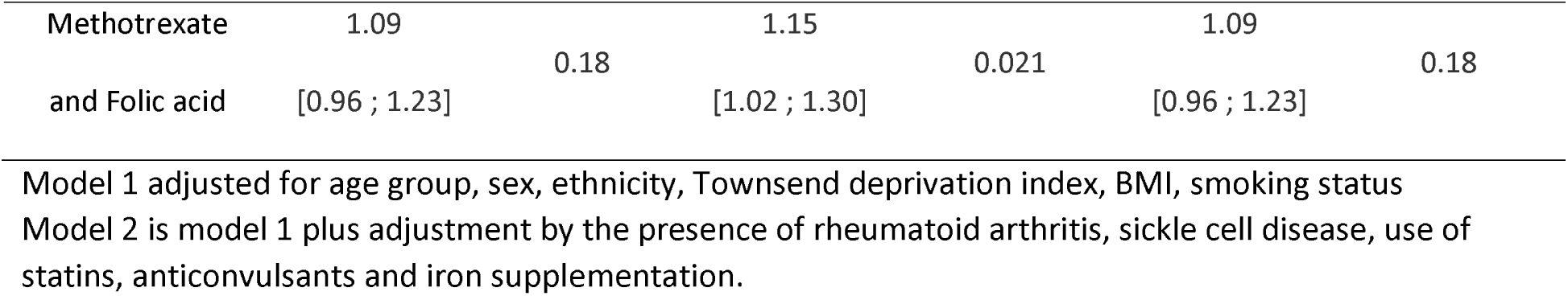
COVID-19 diagnosis in people prescribed methotrexate and / or folic acid in the UKBB,

|  | *Unadjusted* |  | *Model 1* |  | *Model 2* |  |
| --- | --- | --- | --- | --- | --- | --- |
|  | OR | *P* | OR | *P* | OR | *P* |
|  | [95% CI] |  | [95% CI] |  | [95% CI] |  |
| Neither Folic acid |  |  |  |  |  |  |
| nor | 1.0 | - | 1.0 | - | 1.0 | - |
| Methotrexate |  |  |  |  |  |  |
| Folic acid only | 1.58 | <0.001 | 1.60 | <0.001 | 1.51 | <0.001 |
|  | [1.49 ; 1.68 ] |  | [1.50 ; 1.70] |  | [1.42 ; 1.61] |  |
| Methotrexate | 0.94 | 0.83 | 0.89 | 0.72 | 0.86 | 0.64 |
| only | [0.51 ; 1.72] |  | [0.48 ; 1.65] |  | [0.47 ; 1.6] |  |
| Methotrexate | 1.09 |  | 1.15 |  | 1.09 |  |
|  |  | 0.18 |  | 0.021 |  | 0.18 |
| and Folic acid | [0.96 ; 1.23] |  | [1.02 ; 1.30] |  | [0.96 ; 1.23] |  |

Model 1 adjusted for age group, sex, ethnicity, Townsend deprivation index, BMI, smoking status
Model 2 is model 1 plus adjustment by the presence of rheumatoid arthritis, sickle cell disease, use of statins, anticonvulsants and iron supplementation.

### Association with mortality related to a COVID-19 diagnosis

In the general population, compared with people prescribed neither folic acid nor methotrexate, individuals prescribed folic acid had a significant association with mortality related to COVID-19 in Model 1 (OR 2.91 [2.38 ; 3.55]) (Table 3). In Model 2, which included a diagnosis of RA, sickle cell disease, prescription of anticonvulsants, statins and iron supplements, this association was maintained (OR 2.64 [2.15 ; 3.24]). There were no deaths reported in individuals diagnosed with COVID-19 who were prescribed only methotrexate (N = 11). The prescription of methotrexate in combination with folic acid was not associated with an increased odds for death after diagnosis of COVID-19 in Model 1 (Table 3) (1.26 [ 0.70 ; 2.30]) or Model 2 (1.07 [0.57 ; 1.98]). The risk for mortality after COVID-19 diagnosis was of similar magnitude with the prescription of folic acid in both men and women in Model 2 (OR 2.59 [2.00 ; 3.36] and 2.72 [1.93 ; 3.84], respectively) (Table S3). In both men and women co-prescription of methotrexate attenuated the association.

**Table 3.** The association of prescription of methotrexate and folic acid with COVID-19-related death in

|  | *Unadjusted* |  | *Model 1* |  | *Model 2* |  |
| --- | --- | --- | --- | --- | --- | --- |
|  | OR, | *P* | OR, | *P* | OR, | *P* |
|  | [95% CI] |  | [95% CI] |  | [95% CI] |  |
| Neither Folic acid<br>nor Methotrexate | 1.0 | - | 1.0 | - | 1.0 | - |
| Folic acid only | 5.14 | <0.001 | 2.91 | <0.001 | 2.64 | <0.001 |
|  | [4.23 ; 6.24] |  | [2.38 ; 3.55] |  | [2.15 ; 3.24] |  |
| Methotrexate and<br>folic acid | 1.47 | 0.21 | 1.26 | 0.44 | 1.07 | 0.84 |
|  | [0.81 ; 2.67] |  | [0.70 ; 2.30] |  | [0.57 ; 1.98] |  |

the UK Biobank.
Model 1 adjusted for age group, sex, ethnicity, Townsend deprivation index, BMI, smoking status
Model 2 is model 1 plus adjustment by the presence of rheumatoid arthritis, sickle cell disease, use of statins, anticonvulsants and iron supplementation.

To account for improvements in outcome with COVID-19, changes in public health measures and emergence of different SARS-CoV-2 lineages over time ^16^ we tested for association with death in the COVID-19-positive cohort including also a quarterly (3-monthly) categorical time variable for diagnosis of COVID-19 using Model 2 (Table S4). This revealed a similar pattern of association with death - there was association with increased risk of death in those prescribed folic acid only (OR 1.46 [1.16 ; 1.83]) but not in the group prescribed both folic acid and methotrexate (OR 0.96 [0.50 ; 1.83]).

## Discussion

In this population-based analysis, we report 1.5-fold increased risk for COVID-19 diagnosis and 2.6-fold increased risk for COVID-19-related death among those who had been prescribed folic acid supplementation. The prescription of methotrexate was not associated with an increased risk of diagnosis of COVID-19 and we were not able to make an estimate for COVID-19-related death in the small sample of those prescribed methotrexate only. Notably, those prescribed methotrexate and folic acid did not have an increased risk for COVID-19 diagnosis or associated death, indicating that methotrexate might attenuate an increased risk for COVID-19 diagnosis and death conferred by folic acid.

In the context of SARS-Cov-2 infection it is established that hijacking of cellular metabolic pathways is important for viral replication.^17^ Zhang et al described that SARS-CoV-2 remodels host folate and one-carbon metabolism at the post-transcriptional level to support de novo purine synthesis, bypassing viral shutoff of host translation. ^9^ This suggests that viral replication could be sensitive to folate inhibitors, such as methotrexate. Intracellular glucose and folate are depleted in SARS-CoV-2-infected cells, and viral replication is exquisitely sensitive *in vitro* to inhibitors of folate and one carbon metabolism, notably methotrexate.^9^ Stegmman et al, based on cell culture experiments, reported that methotrexate alone or in combination with remdesivir limits the replication of SARS-CoV-2.^18^ With the caveat that our study is observational epidemiology and causality cannot be inferred, our study does support the possibility that external folate supply facilitates the production of large amounts of virus, contributing to clinical infection and mortality. Our study also supports the notion that SARS-CoV-2 replication is enhanced by folate supply by our finding that co-prescription of an antifolate (methotrexate) can ameliorate the possibly adverse effect of supplementation with folic acid on COVID-19 outcomes.

There is also evidence that inadequate folate status may be harmful in the context of host resistance to infection with SARS-CoV-2. In addition to the well-recognized complication of anemia, folate deficiency has other detrimental health effects, including suppression of immune function.^19^ Additional support for the concept that adequate folate status is important in COVID-19 outcomes is provided by the observation that folate deficiency was associated with poorer outcomes in a cohort of COVID-19 patients.^20^ (It is important to note that it is possible that in the study by Itelman et al,^20^ if increased folate levels were causal of COVID-19 diagnosis and poor outcomes, that the association with lower folate levels could have been caused by selection (collider) bias.^21^) Vitamin B12 deficiency has also been proposed as a factor related to poor COVID-19 outcomes, presumed to be through the induction of functional folate deficiency.^22^ A drug-protein structure interaction analysis raises the possibility that folate blocks the 3CL hydrolase enzyme, which may affect viral entry and replication.^23^ It is therefore possible that both inadequate and excessive amounts of folate may be detrimental to host resistance to SARS-CoV-2 infection and that there may be an optimal range of physiological folate status related to host-resistance to COVID-19 infection and severity.

Data from the COVID-19 Global Rheumatology Alliance describes that a number of immunomodulatory drugs used in rheumatology are associated with an increased risk of infection and death compared with methotrexate. ^8^ Being on no DMARD therapy was associated with an increased risk of death with COVID-19 (OR 2.11 [1.48 ; 3.01]), which could be interpreted as either a protective effect of methotrexate or an increased risk for death associated with poor rheumatic disease control. The authors of the study additionally noted that people not on DMARD therapy had increased use of glucocorticoids meaning that confounding by indication cannot be ruled out as an explanation.^24^ Methotrexate was also associated with lower odds for death when compared with sulfasalazine, other immunosuppressants and rituximab. In no case was methotrexate associated with an increased risk for death. The COVID-19 Global Rheumatology Alliance study did not explore the effect of folic acid supplementation in the setting of methotrexate, although it is it is highly likely that almost all patients on methotrexate also were receiving folic acid supplementation. Considering the widespread use of folic acid supplements and proposals to abandon entirely tolerable upper intake levels for folic acid ^25^ it would be prudent to monitor the effect of increased folic acid intake at a population level on COVID-19 morbidity and mortality, particularly at the upper end of folic acid intake.

A number of limitations of our analysis are important to note. One, given the small size of the methotrexate-only group and that there were no deaths related to COVID-19 in this group we could not test a beneficial effect on mortality of methotrexate in isolation. It is uncommon to find patients with methotrexate prescribed without supplemental folic acid as this is the standard of care. Second, over the time period of this study (March 2020 – November 2021), COVID-19 outcomes (i.e. death) will have been influenced by the development of clinical treatments including antiviral drugs and monoclonal antibodies, changes to public health measures and the appearance of new COVID-19 strains ^16^. We were unable to account for these factors in the population-based analysis however we attempted to account for this in the analysis within the COVID-19-positive group by including a time variable (Table S4). Third, findings are not necessarily generalizable outside of the middle-aged (>45 years of age) white European cohort that dominates the UK Biobank. Fourth, the full extent of SARS-CoV-2 infection is not known in the UK population due to incomplete testing rates early in the pandemic. Fifth, prescription data were single script from General Practitioners only, and it was not possible to ascertain compliance or whether participants were taking the prescribed medication during the COVID-19 pandemic although we attempted to account for this by only using prescription information from 2019 and 2020. Sixth, although we included RA in Model 2, we were unable to account for any potential effect of disease activity in RA in people prescribed folic acid. Disease activity negatively impacts death from COVID-19 outcomes.^8^ Finally, while it is a strength of our study that mandatory fortification of the UK diet with folic acid had not been introduced during the period of our study and thus did not confound our analysis, we were unable to account for the lower-dose over the counter folic acid supplementation available in the UK (400 micrograms being the most common formulation for over the counter tablets) because there were no self-report data on the use of folic acid supplementation.

In conclusion, and despite the limitations of our study enumerated above, our data support the hypothesis that increased folate resulting from folic acid prescription could contribute to a higher probability of contracting clinically-detectable infection with SARS-CoV2 and to an increase in the risk of death following the infection. The study population was drawn from the >45 year old segment of the UK population and is predominantly of white European ethnicity, therefore our findings have reduced generalizability to younger people, to other ethnic groups and to other countries. Nevertheless, our findings justify future studies on the influence of folic acid supplementation on COVID-19 outcomes, particularly in pregnant women and people on anticonvulsants requiring supplementary folic acid.

As a final comment, we point out that attention is currently being directed toward establishing whether excessive intake of folate, particularly in the form of folic acid, may have undesirable and potentially deleterious effects.^26^ The possibility that susceptibility to COVID-19 infection and its serious and even fatal complications may be affected by folic acid intake and folate status should be thoroughly investigated.

## Data Availability

All summary data produced in the present study are available upon reasonable request to the authors. The authors are unable to share the individual level data from the UK Biobank.

## Acknowledgements

This research was conducted using the UK Biobank Resource under Application Number 12611. We sincerely thank all participants.

## Author contributions

All authors substantially contributed to study conception and design, to acquisition and analysis of data and interpretation of results. All authors contributed to drafting the article and critical revision and all authors approved the final version. RKT and TRM directly accessed and verified the underlying data reported in the manuscript.

## Data sharing

All data utilised in this study were accessed from the publicly available UK Biobank Resource under Application Number 12611. These data cannot be shared with other investigators.

## Figure legends

**Table S1.** Medications co-prescribed with folic acid on >5% of occasions

| Medication/Supplement | Number of prescriptions* | Percentage of folic acid prescriptions |
| --- | --- | --- |
| Folic acid | 177,918 | 100.00 |
| Methotrexate | 45,697 | 25.68 |
| Atorvastatin | 41,296 | 23.21 |
| Lansoprazole | 35,611 | 20.02 |
| Omeprazole | 32,909 | 18.50 |
| Bisoprolol | 26,445 | 14.86 |
| Levothyroxine | 25,482 | 14.32 |
| Ramipril | 21,990 | 12.36 |
| Aspirin | 21,696 | 12.19 |
| Paracetamol | 20,830 | 11.71 |
| Amlodipine | 18,354 | 10.32 |
| Simvastatin | 16,878 | 9.49 |
| Amitriptyline | 13,573 | 7.63 |
| Furosemide | 12,983 | 7.30 |
| Metformin | 11,460 | 6.44 |
| Hydroxychloroquine | 11,038 | 6.20 |
| Clopidogrel | 10,918 | 6.14 |
| Prednisolone | 10,475 | 5.89 |
| Alendronic acid | 10,198 | 5.73 |

\*General practice prescription data from 1st of January 2019 through to 27<sup>th</sup> September 2021

**Table S2.** Sex-specific associations between prescription of methotrexate, folic acid, and the combination of both with the risk of COVID-19

|  | COVID-19 diagnosis - Male |  |  |  |  |  | COVID-19 diagnosis -Female |  |  |  |  |  |
| --- | --- | --- | --- | --- | --- | --- | --- | --- | --- | --- | --- | --- |
|  | *Unadjusted* |  | *Model 1* |  | *Model 2* |  | *Unadjusted* |  | *Model 1* |  | *Model 2* |  |
|  | OR,<br>[95% CI] | *P* | OR,<br>[95% CI] | *P* | OR,<br>[95% CI] | OR,<br>[95% CI] | OR,<br>[95% CI] | *P* | OR,<br>[95% CI] | *P* | OR,<br>[95% CI] | *P* |
| Neither folic acid<br>nor methotrexate | 1.0 | - | 1.0 | - | 1.0 | - | 1.0 | - | 1.0 | - | 1.0 | - |
| Folic acid only | 1.52<br>[ 1.39 ;<br>1.66 ] | <0.001 | 1.57<br>[ 1.44 ;<br>1.72 ] | <0.001 | 1.50<br>[ 1.37 ;<br>1.64 ] | <0.001 | 1.64<br>[ 1.51 ;<br>1.77 ] | <0.001 | 1.61<br>[ 1.48 ;<br>1.75 ] | <0.001 | 1.52<br>[ 1.39 ;<br>1.65 ] | <0.001 |
| Methotrexate<br>only | 0.96<br>[ 0.39 ;<br>2.38 ] | 0.93 | 0.93<br>[ 0.37 ;<br>2.32 ] | 0.88 | 0.92<br>[ 0.37 ;<br>2.30 ] | 0.86 | 0.92<br>[ 0.4 ;<br>2.11 ] | 0.85 | 0.87<br>[ 0.38 ;<br>1.99 ] | 0.74 | 0.83<br>[ 0.36 ;<br>1.90 ] | 0.65 |
| Methotrexate<br>and folic acid | 1.08<br>[ 0.89 ;<br>1.31 ] | 0.45 | 1.15<br>[ 0.94 ;<br>1.40 ] | 0.17 | 1.11<br>[ 0.91 ;<br>1.36 ] | 0.29 | 1.11<br>[ 0.95 ;<br>1.29 ] | 0.19 | 1.16<br>[ 0.99 ;<br>1.36 ] | 0.060 | 1.07<br>[ 0.91 ;<br>1.26 ] | 0.38 |

diagnosis
Model 1 adjusted for age group, sex, ethnicity, Townsend deprivation index, BMI, smoking status
Model 2 is model 1 plus adjustment by the presence of rheumatoid arthritis, sickle cell disease, use of statins, anticonvulsants and iron supplementation.

**Table S3.** Sex-specific associations between prescription of methotrexate, folic acid, and the combination of both with the risk of COVID-19

|  | COVID-19 related death - Male |  |  |  |  |  | COVID-19 related death - Female |  |  |  |  |  |
| --- | --- | --- | --- | --- | --- | --- | --- | --- | --- | --- | --- | --- |
|  | *Unadjusted* |  | *Model 1* |  | *Model 2* |  | *Unadjusted* |  | *Model 1* |  | *Model 2* |  |
|  | OR,<br>[95%<br>CI] | *P* | OR,<br>[95%<br>CI] | *P* | OR,<br>[95%<br>CI] | OR, [95%<br>CI] | OR,<br>[95%<br>CI] | *P* | OR,<br>[95%<br>CI] | *P* | OR,<br>[95%<br>CI] | *P* |
| Neither folic acid nor methotrexate | 1.0 | - | 1.0 | - | 1.0 | - | 1.0 | - | 1.0 | - | 1.0 | - |
| Folic acid only | 4.73<br>[3.71;<br>6.04] | <0.001 | 2.83<br>[2.20;<br>3.64 ] | <0.001 | 2.59<br>[2.00;<br>3.36] | <0.001 | 5.67<br>[4.10;<br>7.82 ] | <0.001 | 3.07<br>[2.20;<br>4.28] | <0.001 | 2.72<br>[1.93;<br>3.84] | <0.001 |
| Methotrexate and folic acid | 2.00<br>[0.99;<br>4.03] | 0.053 | 1.62<br>[0.80;<br>3.26] | 0.18 | 1.54<br>[0.75;<br>3.15] | 0.24 | 1.01<br>[0.32;<br>3.16] | 0.99 | 0.80<br>[0.25;<br>2.50] | 0.70 | 0.56<br>[0.17;<br>1.84] | 0.34 |

related mortality.
Model 1 adjusted for age group, sex, ethnicity, Townsend deprivation index, BMI, smoking status
Model 2 is model 1 plus adjustment by the presence of rheumatoid arthritis, sickle cell disease, use of statins, anticonvulsants and iron supplementation.

**Table S4.** Risk of death related to COVID-19 in the COVID-19-positive group

|  | *Unadjusted* |  | *Model 1* |  | *Model 2* |  |
| --- | --- | --- | --- | --- | --- | --- |
|  | OR<br>[95% CI] | *P* | OR<br>[95% CI] | *P* | OR<br>[95% CI] | *P* |
| Neither folic acid nor methotrexate | 1.0 | - | 1.0 | - | 1.0 | - |
| Folic acid only | 3.58 [ 2.93 ;<br>4.39 ] | <0.001 | 1.82 [ 1.47 ;<br>2.26 ] | <0.001 | 1.46 [ 1.16 ;<br>1.83 ] | 0.0011 |
| Methotrexate and folic acid | 1.37 [ 0.75 ;<br>2.52 ] | 0.31 | 1.03 [ 0.55 ;<br>1.91 ] | 0.94 | 0.96 [ 0.50 ;<br>1.83 ] | 0.90 |

Model 2 additionally adjusted by quarterly categorical time variable.
A total of 25,816 COVID-19-positive cases were analyzed. 217 COVID-19-positive cases who were diagnosed after 13<sup>th</sup> September 2021 (28 days before the last recorded death) were removed from the cohort given the unknown outcome in these individuals.

